# Statewide Impact of COVID-19 on Social Determinants of Health - A First Look: Findings from the Survey of the Health of Wisconsin

**DOI:** 10.1101/2021.02.18.21252017

**Authors:** Kristen M.C. Malecki, Amy A. Schultz, Maria Nikodemova, Matt C. Walsh, Andrew J. Bersch, Jacquie Cronin, Lisa Cadmus-Bertram, Corinne D. Engelman, Julia R. Lubsen, Paul E. Peppard, Ajay K. Sethi

**Author notes:** Corresponding Author: Kristen MC Malecki PhD, MPH, Associate Professor, Department of Population Health Sciences, 610 N. Walnut Street, WARF 605, Madison, WI 53726.

## Abstract

There is an urgent need to track the early and ongoing impact of the COVID-19 pandemic on population health from local to global scales. At the same time, there is an overall lack of U.S. state-specific surveillance data tracking social determinants of health (SDOH) and associations with population well-being, individual mitigation and coping strategies, family dynamics and other economic shocks of the pandemic in populations. Statewide data can offer important insights into how SDOH shape the long-term effects of COVID-19 in the population since implementation of many policies and programs varied widely early on in the pandemic. In May of 2020, the Survey of the Health of Wisconsin (SHOW) program launched a statewide online/phone survey of early and ongoing impacts of COVID-19 on health and well-being across diverse communities and families. The goal of this study is to provide descriptive data including perceived COVID-19 risks, access to and results of COVID-19 antigen testing, individual mitigation and coping strategies, family dynamics and other economic shocks of the pandemic on health and mental health in populations. Key findings include higher rates of testing and perceived past infection from COVID-19 among non-white respondents. Higher economic shifts and job changes in female vs male respondents. Families with children reported overall higher levels of stress, and stress from the pandemic. There were urban and rural differences in changes to access to care. Rural regions, which had a lower prevalence of infections early in the pandemic as compared to urban areas, also reported fewer delays or missed appointments due to COVID-19. Key findings show that SDOH are shaping impacts of health and well-being early on in the pandemic and future longitudinal follow-up will be important to shape policies and programs well into the future.

## INTRODUCTION

The SARS-COV-2 pandemic, which began sweeping the United States in early 2020, has impacted morbidity and mortality across the globe and resulted in shocks to economics, social structure and daily living with long-term consequences. The acute need to focus on disease incidence and spread in communities was followed by the need to examine the changing social determinants of health (SDOH). Key to the definition of SDOH is that several upstream factors have significant impacts on population health and well-being including social and community context, access to and quality of health care, economic stability, access to and quality of education, and the physical environments in which people live (U.S. DHHS, 2020). Each of these macro level factors also influence the resources and assets available within communities to respond to and address the more downstream economic and mental health consequences of COVID-19. Pandemics have the potential to shift social determinants not only among the most vulnerable, lower income communities, but among the broader general population. There is an urgent need to track the early and ongoing impact of the COVID-19 pandemic on population health from local to global scales. At the same time, there is an overall lack of U.S. state-specific surveillance data tracking SDOH and associations with population well-being, individual mitigation and coping strategies, family dynamics and other economic shocks of the pandemic in populations.

In May of 2020, the Survey of the Health of Wisconsin (SHOW) program launched a statewide online/phone survey of early and ongoing impacts of COVID-19 on health and well-being across diverse communities and families. The SHOW program, originally launched in 2008, maintains a large state-wide representative cohort of Wisconsin residents. Wisconsin is a geographically diverse state with over one-third of the population living in remote, rural areas and a substantial urban population living in several metropolitan areas with unique cultural, political and social contexts. Milwaukee, the largest Wisconsin city, is a racially and ethnically diverse city with a large urban core of Black and LatinX populations. Wisconsin also shares many similarities with other Midwestern states and offers an important representation of life in the north-central region of the United States.

At the time of this baseline survey, COVID-19 prevalence was relatively low and varied regionally across the state of Wisconsin. The Wisconsin general population is about one-third rural, one third urban and one-third suburban. There was wide variation in local policy response and implementation of COVID-19 mitigation strategies across the seventy-two counties. The largest metropolitan areas in Southeastern WI, (Milwaukee, Racine and Kenosha) had the highest incidence of COVID-19 infections early in the pandemic, while incidence in rural regions rose later. Results presented here offer a first glimpse at population health during this early period of the COVID-19 pandemic among these diverse Wisconsin populations. We examined how SDOH including gender, urbanicity, education, race and family structure was influence COVID-19 testing and mitigation, perception, economic impacts, changes in family dynamics, access to care, coping strategies and overall mental health and well-being.

This study provides descriptive data from the first wave of the SHOW COVID-19 impact survey (May-June, 2020) of the pandemic. Statewide data can offer important insights into how SDOH shape the long-term effects of COVID-19 in the population because there is implementation of many policies and programs varied widely early on in the pandemic. An overview of survey methodologies and a summary of key findings from this early period in the pandemic by select domains stratified across key SDOH – urbanicity, economic, education and gender – is presented.

## METHODS

### Study Participants and Recruitment

The COVID-19 Impact Survey was conducted as part of the ongoing Survey of the Health of Wisconsin (SHOW) program. SHOW is a population-based health examination survey designed to collect objective and subjective data on health and on a broad number of social health determinants described in detail elsewhere (Nieto, et al. 2010). This first wave of COVID-19 impact survey sample included all past adult SHOW participants from 2008 through 2020 who consented to be contacted for future research and who were not deceased as of the first day of recruitment and it was restricted to those who provided a valid email address or phone number for future contacts. See supplementary Fig 1 for a description of the study sample. The online survey was conducted between May 18-July 5, 2020 by emailing a unique link to each participant. Participants without a valid email address were contacted by phone to request their email. If participants were reachable by phone but did not have an email address or were unable to complete the survey online, then they were offered a shortened version of the survey that was administered over the phone within the same time. The study was approved by the University of Wisconsin-Madison Health Science Institutional Review Board. As the survey was launched, COVID-19 restrictions and safety precautions precluded SHOW study staff from working in secure office settings to print, prepare, and mail paper invite letters and postcards to those without email addresses, those who were not accessible by phone or those who did not have internet access. However, towards the end of the participation window (mid-June 2020), staff were able to mail postcards to hard-to-reach participants and to update contact information for the future COVID-19 survey waves and other study opportunities. All participants who completed the online COVID-19 or telephone survey received a $25 electronic gift card.

### COVID-19 Impact Survey Instrument

The survey consists of 9 domains including (1) COVID-19 perceptions, beliefs, behaviors; (2) economic well-being; (3) food security, diet and housing; (4) personal, social and community context; (5) health and healthcare access; (6) mental and emotional health; (7) information sources and literacy; (8) lifestyle behaviors; and (9) caregiving (Table 1). The majority of questions were multiple choice or formatted as a Likert scale. At the end of the survey participants were asked to consent for future linkage with electronic health records. Renumeration was either emailed or mailed to participants in the form of an electronic or physical gift-card. A complete copy of the survey instrument is available in supplemental materials.

**Table 1.** SHOW’s COVID-19 Community Impact Survey Domains

|  |  |
| --- | --- |
| **COVID-19 – Perceptions, Beliefs, Behaviors** | **Health & Healthcare Access** |
| *History of exposure & testing* | *Chronic health conditions* |
| *Symptoms & hospitalization* | *Flu vaccine history* |
| *Mitigation perceptions & behaviors* | *Medication / treatment access* |
| *Perceived threat* | *Missed appointments* |
| *Coping strategies* | *Delayed procedures/surgeries* |
| *Vaccine willingness* | *Advance care planning* |
| *Lasting symptoms* | *Reproductive health* |
| **Economic Well Being** | **Mental & Emotional health** |
| *Employment status & job type* | *Anxiety and Stress screener* |
| *Changes in employment* | *Previous Mental Health Diagnosis* |
| *Insurance status & type* | *PTSD screener* |
| *Change in insurance* | *Emotional support* |
| *Retirement funds loss* | *COVID-19 specific stressors* |
| **Food security, diet and housing** | **Information Sources and Literacy** |
| *Use of benefit programs before/after* | *Trusted news sources* |
| *Worried about food lasting* | *Internet access* |
| *Changes in eating habits / meals* | *Know how to find info on internet* |
| *Foods consumed more/less* | **Lifestyle behaviors** |
| *Unable to pay rent/mortgage* | *Sleep quality* |
| *Had to relocate* | *Physical activity* |
| **Personal, social & community context** | *Alcohol consumption* |
| *Resilience* | *Smoking habits* |
| *Social cohesion* | **Caregiving** |
| *Risk Taking, trust, altruism* | *Children /Adults* |
| *Discrimination* | *Stress, strain & coping* |
| *Political Empowerment* | *Family activities* |

### Data Analyses

The data presented are key indicators within a few of the survey domains. Descriptive analyses include frequencies and proportions of group-levels responses and stratification by demographic factors (gender, education, urbanicity). All analyses were performed with SAS v9.4.

## RESULTS

### Survey participants

Participation was well distributed geographically, having proportional representation from urban and rural areas. No differences of gender, education, self-reported health, asthma, COPD, or cardiovascular disease were observed when comparing urban and rural participants (Table 2). Participants residing in rural areas were more likely to be in the older (>60 years), white, or have diabetes compared to those in urban areas.

**Table 2.** Demographic and general characteristics of participants by urbanicity

|  | Survey Sample<br>(n = 1378)<br>% | Urban<br>(n=945)<br>% | Rural<br>(n=433)<br>% | P-trend |
| --- | --- | --- | --- | --- |
| **Gender** |  |  |  | 0.4736 |
| Male | 36.5 | 35.9 | 37.9 |  |
| Female | 63.5 | 64.1 | 62.1 |  |
| **Current Age (in years)** |  |  |  | <.0001 |
| 18-39 | 18.7 | 22.4 | 10.4 |  |
| 40-59 | 35.3 | 36.0 | 33.7 |  |
| 60-94 | 46.1 | 41.6 | 55.9 |  |
| **Race** |  |  |  | <.0001 |
| White (non-Hispanic) | 86.7 | 82.7 | 95.4 |  |
| AA (non-Hispanic) | 7.6 | 11.0 | 0.0 |  |
| Hispanic | 2.3 | 2.8 | 1.4 |  |
| Other (non-Hispanic) | 3.4 | 3.5 | 3.2 |  |
| **Education** |  |  |  | 0.6753 |
| H.S./GED or less | 2.3 | 2.2 | 2.3 |  |
| Some college | 48.8 | 48.0 | 50.6 |  |
| Bachelors or higher | 48.9 | 49.7 | 47.1 |  |
| **Household Income** |  |  |  | 0.0388 |
| < \$25,000 | 12.8 | 14.4 | 9.2 | |
| \$25,000 - \$49,999 | 22.2 | 22.0 | 22.6 | |
| \$50,000 - \$99,999 | 35.5 | 33.9 | 38.9 | |
| >\$99,999 | 29.5 | 29.6 | 29.3 | |
| **Household size** |  |  |  | 0.0006 |
| 1 | 15.2 | 17.2 | 10.9 |  |
| 2 | 46.1 | 43.5 | 51.6 |  |
| 3-4 | 29.5 | 31.0 | 26.1 |  |
| 5+ | 9.3 | 8.3 | 11.4 |  |
| **Self-reported Health** |  |  |  | 0.2663 |
| Excellent | 11.9 | 12.2 | 11.2 |  |
| Very Good | 47.1 | 45.5 | 50.4 |  |
| Good | 32.6 | 32.8 | 32.1 |  |
| Fair | 7.3 | 8.3 | 5.4 |  |
| Poor | 1.1 | 1.2 | 0.9 |  |
| **Chronic Health Condition** |  |  |  |  |
| Asthma and/or COPD | 11.5 | 12.4 | 9.5 | 0.1138 |
| Cardiovascular disease | 34.4 | 34.0 | 35.3 | 0.6201 |
| Diabetes (any type) | 8.2 | 7.4 | 9.9 | 0.1130 |

Abbreviations: AA: African American; N: number; H.S.: high school; GED: General Education Development test; COPD: Chronic Obstructive Pulmonary Disease Column percents shown.
p-trend: statistically significant difference of demographics by urbanicity using Chi-square test
Urbanicity: considered rural if residence located in rural census block group defined by the U.S. Census Bureau as having fewer than 2,500 people
Cardiovascular disease: if reported history of congestive heart failure, myocardial infarction, angina pectoris, stroke, high cholesterol/hyperlipidemia, or transient ischemic attack

Among the n=5510 eligible adults, 25% (n=1377) completed the online survey, Response Rate 1, and 25.5% answered at least one survey item (n=1403), Response Rate 2. Response rates 1 and 2 were calculated as defined by the American Association for Public Opinion Research (2016). An additional 55 participants completed the shortened phone interview survey (Suppl Fig 1). The participants who completed the survey online were more likely to be female, non-Hispanic white, have a bachelor’s or advanced degree and a higher household income, when compared to non-respondents. Respondents were also more likely to self-report their health as ‘excellent’ and ‘very good’. Chronic health conditions asthma, COPD, and cardiovascular disease did not differ between those who participated and those who did not. However, participants were less likely to have diabetes of any type.

### COVID-19 exposure and testing by race

Irrespective of testing, as of May-June 2020, approximately 11% of participants believed they may have had COVID-19 in the past. Only 6.8% of the sample reported having had a diagnostic COVID-19 test and 4.3% reported a positive test. Only 1% of the sample was told by a healthcare provided that they had COVID-19 (Suppl Table 2). When compared to Whites, Non-whites were 2-times more likely to believe they had COVID (20% vs. 10%), to have been tested for COVID-19 (14% vs. 6%), and 3-times more likely to have been contacted by a healthcare professional about potential exposure to COVID-19 (6% vs. 2%) (Fig 1). Five percent of Non-whites tried to get tested but were turned away, compared with only 2% of Whites. Non-whites were also three times as likely to report being embarrassed to disclose COVID-19 diagnosis to employers or friends (6% vs. 2%), and twice as likely to have experienced stigma or discrimination due to COVID-19.

**Figure 1.**
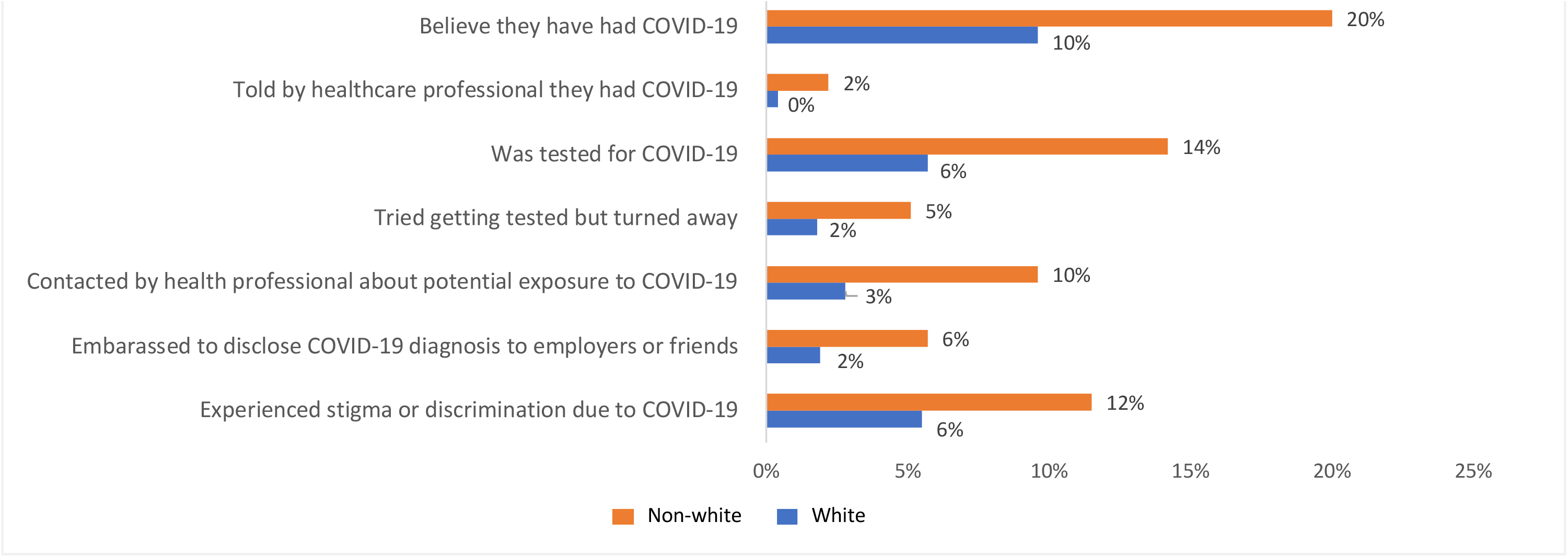
COVID-19 exposure and testing by race. The percent of participants who self-reported the following COVID-19 experiences. The following items represent several individual survey items. Due to voluntary refusal of survey items, missingness varies. See Supplemental Table 2 for sample sizes for each survey item.

### Perceived Efficacy and Behaviors related to COVID-19 Exposure Risks by Education

Perceived effectiveness of different mitigation strategies was assessed and compared by education status. Based on responses to the question “how effective are the following actions for keeping you safe from COVID-19?”, over 90% of respondents believed social distancing, avoiding public spaces & gatherings, and washing hands were somewhat or very effective and 84% reported mask wearing to be effective (Suppl Table 3 and Fig 2). A slightly higher percentage of those with more education reported social distance and washing hands as effective compared to those with less than a bachelor’s degree. Those with less than a college degree were more like to believe praying, avoiding outside exercise, and doing nothing were somewhat or very effective strategies to avoid COVID-19 infection when compared with those with a college degree or higher (57% vs. 39%, 23% vs. 15%, and 21% vs. 12%, respectively).

**Figure 2.**
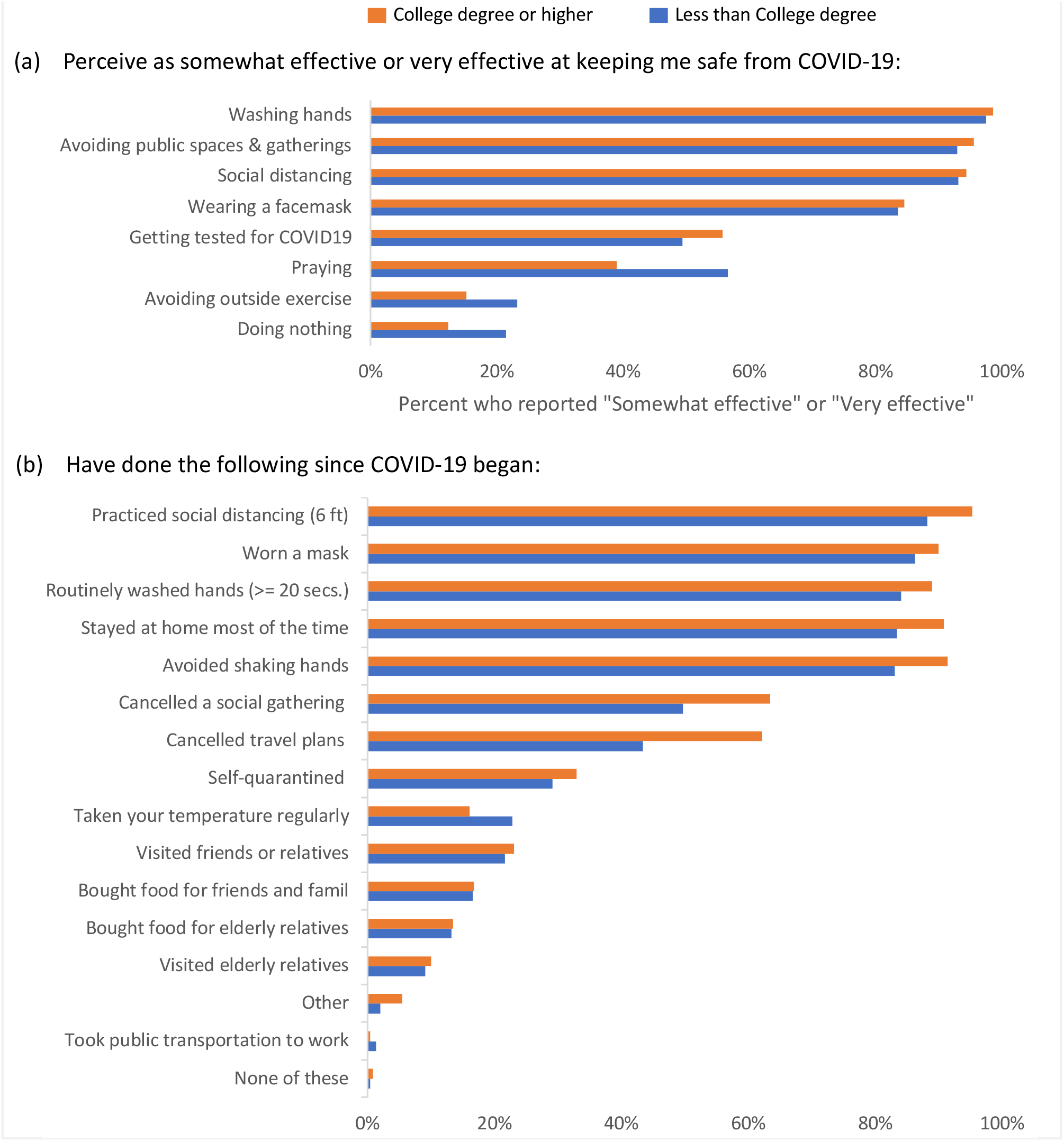
Perceptions and behaviors related to COVID-19 by education level. (a) Percentage of respondents who perceive the following as “Somewhat effective or “Very effective” at keeping them safe from COVID-19. (b) Percentage of respondents who have done the following mitigation behaviors.

**Figure 3.**
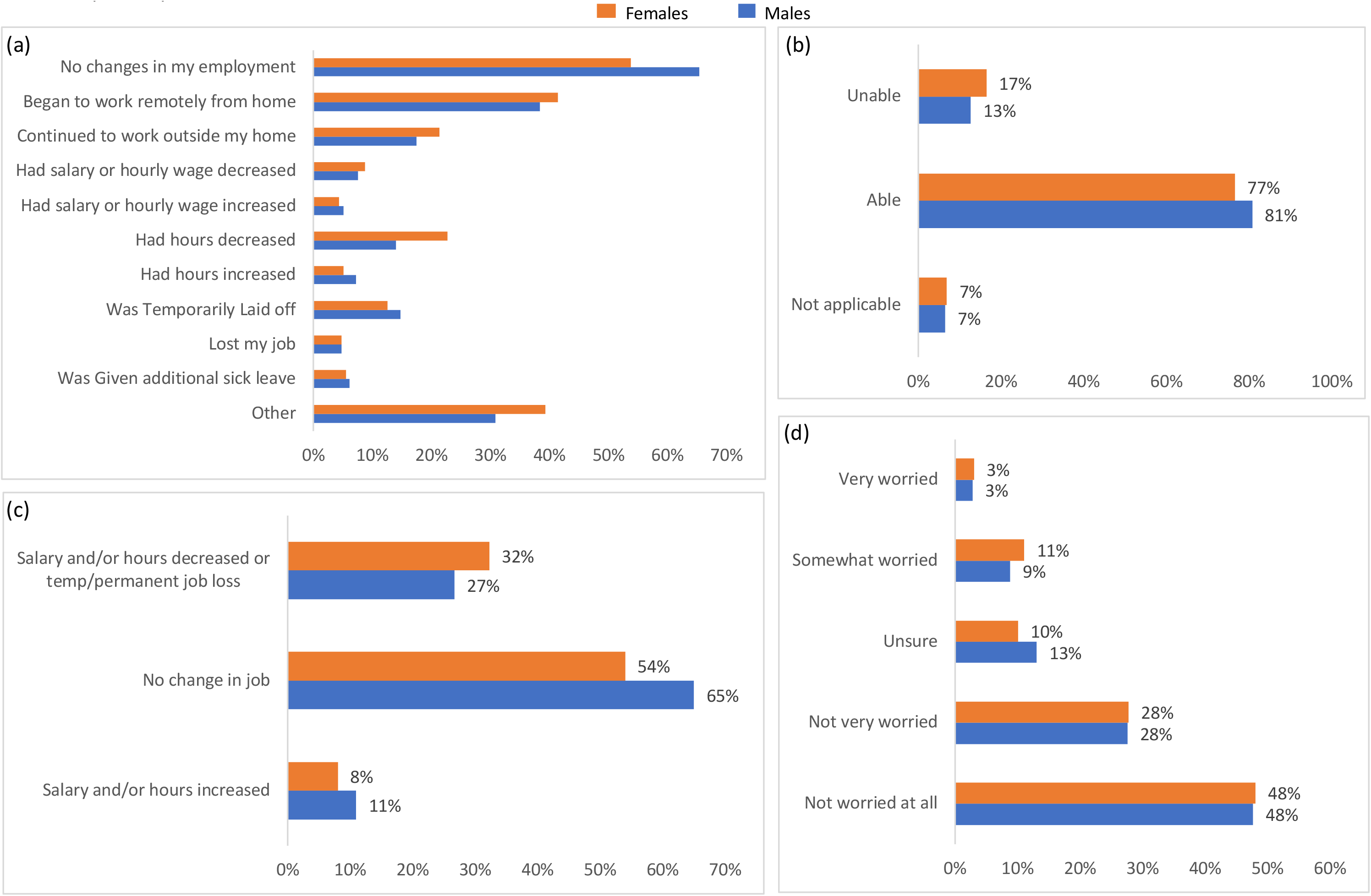
The impact of COVID-19 on employment by gender. (a) Among those who were working prior to COVID-19, their employment experience(s) due to COVID-19 (n=816). (b) Ability to pay rent/mortgage due to COVID-19 (n=812). (c) Derived from figure (a), employment experience due to COVID-19 (n=816). (d) How worried participant is that they will lose their job in the next 3 months (n=780).

A number of common daily activities related to minimizing risk of COVID-19 in the early months of the pandemic were also examined. Overall, less than 25% of the respondents reported visiting elderly individuals, while the majority of participants (> 80%) reported some cancellations of social gatherings, travel plans, avoidance of personal interaction with others and mask wearing during the early stage of the pandemic. Those with a college degree were 10% more likely to cancel a social gathering and 20% more likely to cancel travel plans, but only 4% more likely to have self-quarantined when compared with those with less than a college degree. On the contrary, participants with lower education were more likely to take their temperature regularly (23% vs. 16%). Use of public transit was slightly higher among those without a college education (1% vs. 0%) compared with college educated participants.

### Impacts on Economic Stability due to COVID-19 by Gender

Among those reporting being employed in May/June 2020, approximately 42% of respondents indicated some change in work due to COVID-19 (Supp Table 4). Change in work was defined broadly as a change in office to remote work, furlough or layoff, reduction or increase in hours and/or pay and given additional sick leave. While both men and women were equally concerned about job loss in the next three months, the loss of both work and pay were higher among women compared to men. Women were more likely to experience a job change due to COVID-19 (46%) compared to men (35%). Approximately 11% of men reported an increase in salary and/or hours compared to 8% of women. On the contrary, females were more likely to report a decrease in salary and/or hours due to COVID-19 (32% vs. 27%). Females were also more likely than males to report being unable to pay rent or mortgage because of COVID-19 (17% vs. 13%).

### Healthcare Access and Delays in Care by Urbanicity

Just over half of the participants (56%) did not experience a delay in getting healthcare due to COVID-19, from the start of the pandemic to May/June of 2020 (Fig 4A and Suppl Table 5.) The most frequently reported reason for delay in getting healthcare was due to an appointment being postponed or cancelled due to COVID-19(34%), followed by being afraid to get care because of COVID-19 (8.9%), could not get an appointment soon enough (5.1%), clinic/office was not open (2.4%), and could not get through on the phone (1.2%).

**Figure 4.**
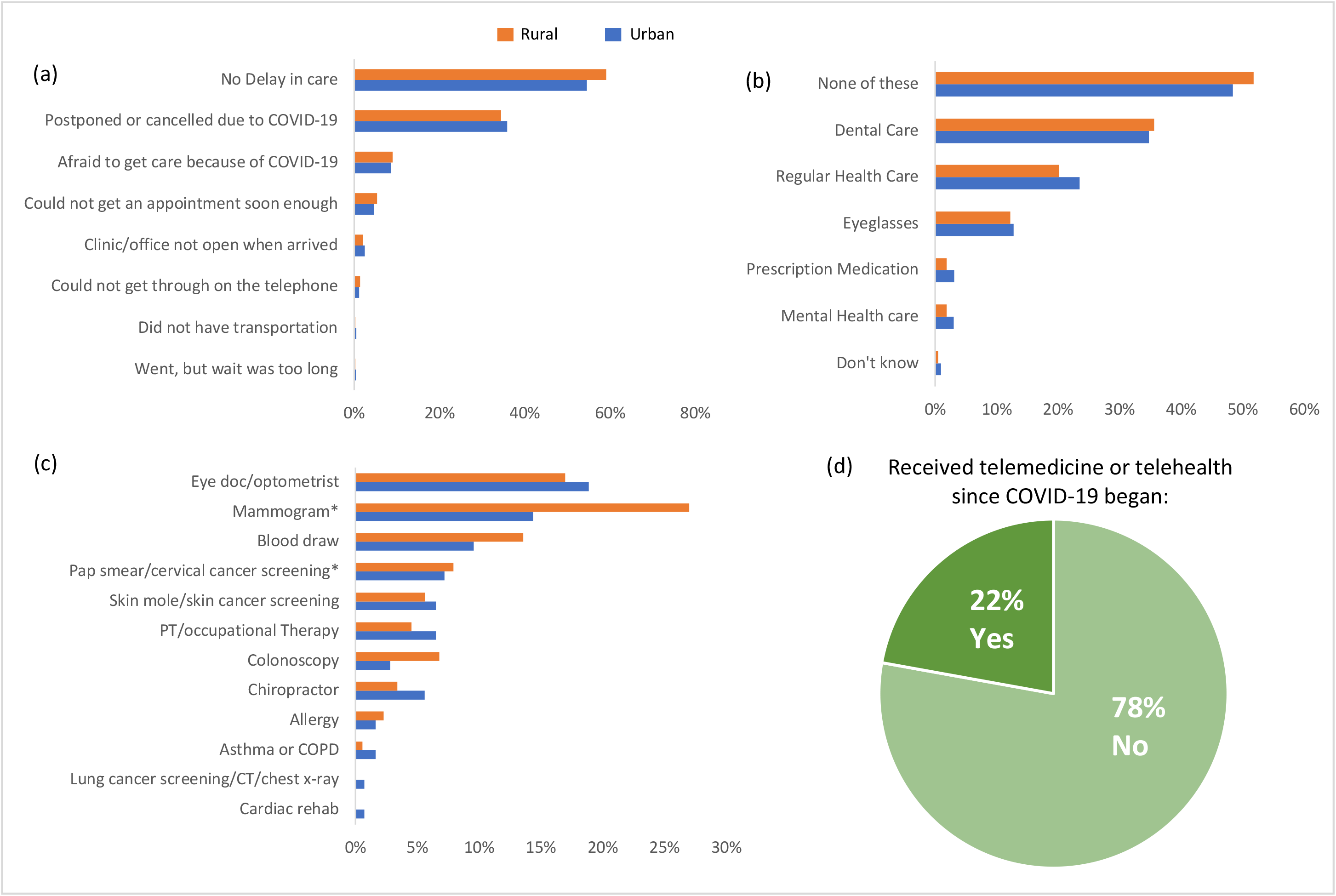
The impact of COVID-19 on receiving health care by urbanicity. (a) Percent who delayed getting care for the following reasons (n= 1378), (b) Percent who indicated there was a time when they needed the following but could not get it because of COVID-19 (n=1378), (c) Among those who experienced delay in care (n=603), the percent who reported the type of appointments and/or procedures missed. (d) The percent who received telemedicine or telehealth since COVDI-19 began (n=1378). Participant is considered to reside in rural location if residence is located in rural census block group defined by the U.S. Census Bureau as having fewer than 2,500 people.

Regular health care, dental care, and eyeglasses (Fig 4B) were among the top three things participants said they could not get because of COVID-19 (22%, 35%, and 13%, respectively). Only slight differences were seen by urbanicity, with rural residents reporting not getting dental care more often and urban residents reporting not getting regular health care more often. About 22% received telemedicine or telehealth since COVID-19 began, with no differences by urbanicity (Fig 4D).

Among those who reported experiencing a delay in healthcare, the most frequently missed appointments reported were eye doctor (18%), mammogram (18%), blood draw (11%), and pap smear/cervical cancer screening appointments (7%) (Fig 4C). Few differences were seen by urbanicity in terms of the type of appointment missed, except for mammogram and blood appointments. Twenty-seven percent of rural female residents who reported a delay in care, missed a mammogram appointment, compared to 14% of urban females who reported a delay in care. Four percent more rural residents reported missing a blood draw when compared to urban residents.

### Changes in Health-Related Behaviors and Coping Mechanisms by Gender

The impact of COVID-19 on health-related behaviors varied by gender in the early stages of the pandemic (Fig 5, Suppl Table 6). The pandemic affected females’ physical activity and alcohol consumption to a greater degree when compared to males. Males were more likely to report the same amount of physical activity compared to before COVID-19 (41% vs 29%) and alcohol consumption (65% vs. 59%), whereas, females were more likely to report being more or less active and drinking more or less now, when compared to before COVID-19. Similarly, when asked what activities survey respondents are doing to cope with stress from COVID-19, females were more likely to report doing more activities, from watching tv, to gardening, reading, playing games, exercising and playing music, than males.

**Figure 5.**
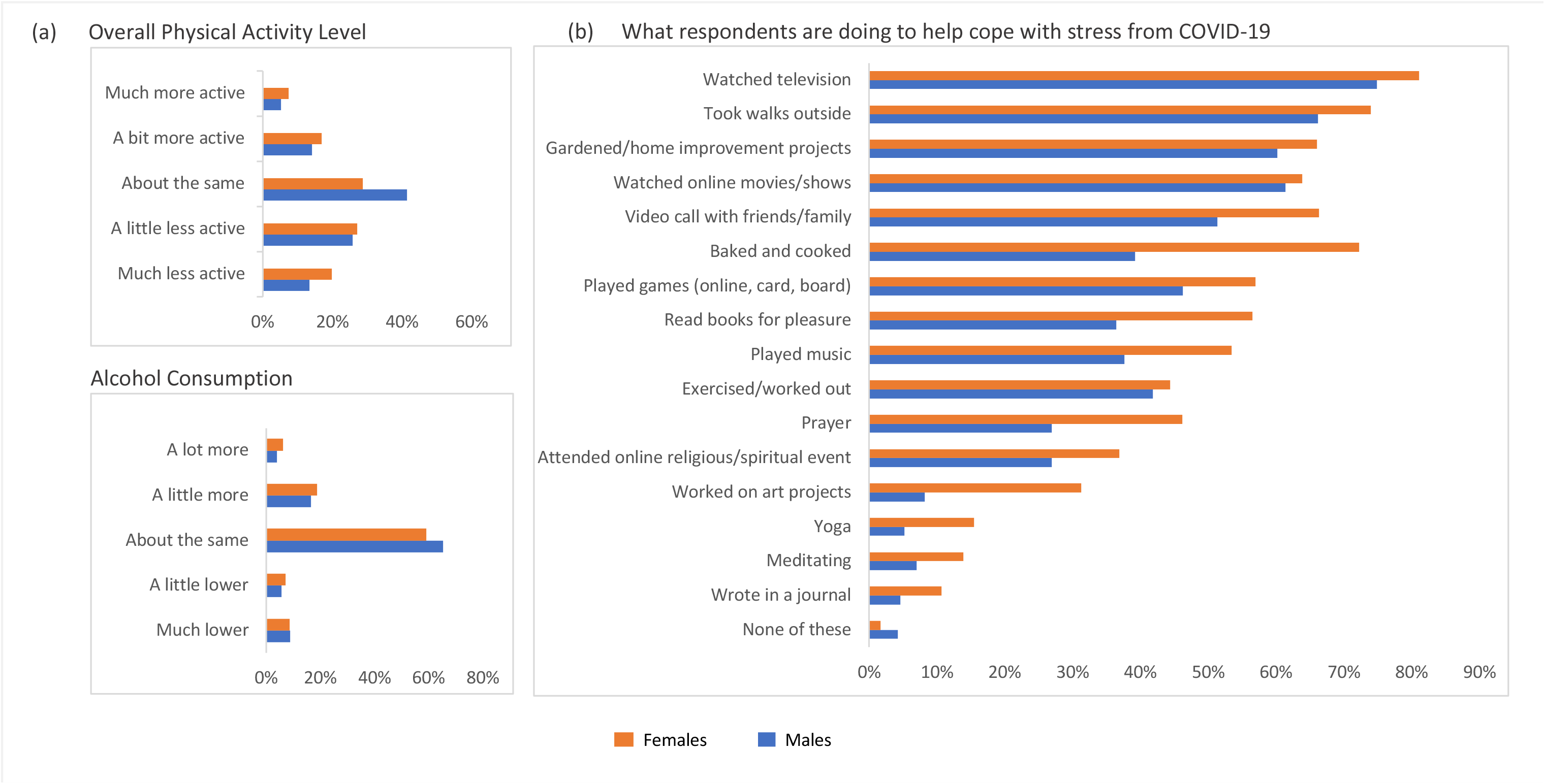
The impact of COVID-19 on health-related behaviors by gender. (a) Respondents reported the degree to which their physical activity level (n=1367) and alcohol consumption (n=1285) changed since COVID-19 began, compared to before COVID-19. Only current alcohol consumers were asked about changes in their alcohol consumption quantity. (b) Percentage of respondents who reported doing the following activities to help cope with stress from COVID-19 (n=1369). (a-b) n=5 nonconforming/transgender were excluded because too few to look at in these analyses.

### Stress, Anxiety and Mental Well-Being by Household Composition

Stark differences in experiencing stress were seen among those with children in the home when compared to those without children in the home (Fig 6). About 30% of survey respondents reported having at least one child (<18 years of age) living in their home (Suppl Table 7a). Among those with children in the home (n=404), 88% reported caring for children in their home during the pandemic. About 6% of the survey respondents care for an adult who has an illness or disability in the home. Adults with children in the home were twice as likely to report moderate-high levels of stress in their job and in caring for others, although adults with children reported higher levels of stress in all domains when compared with adults without children.

**Figure 6.**
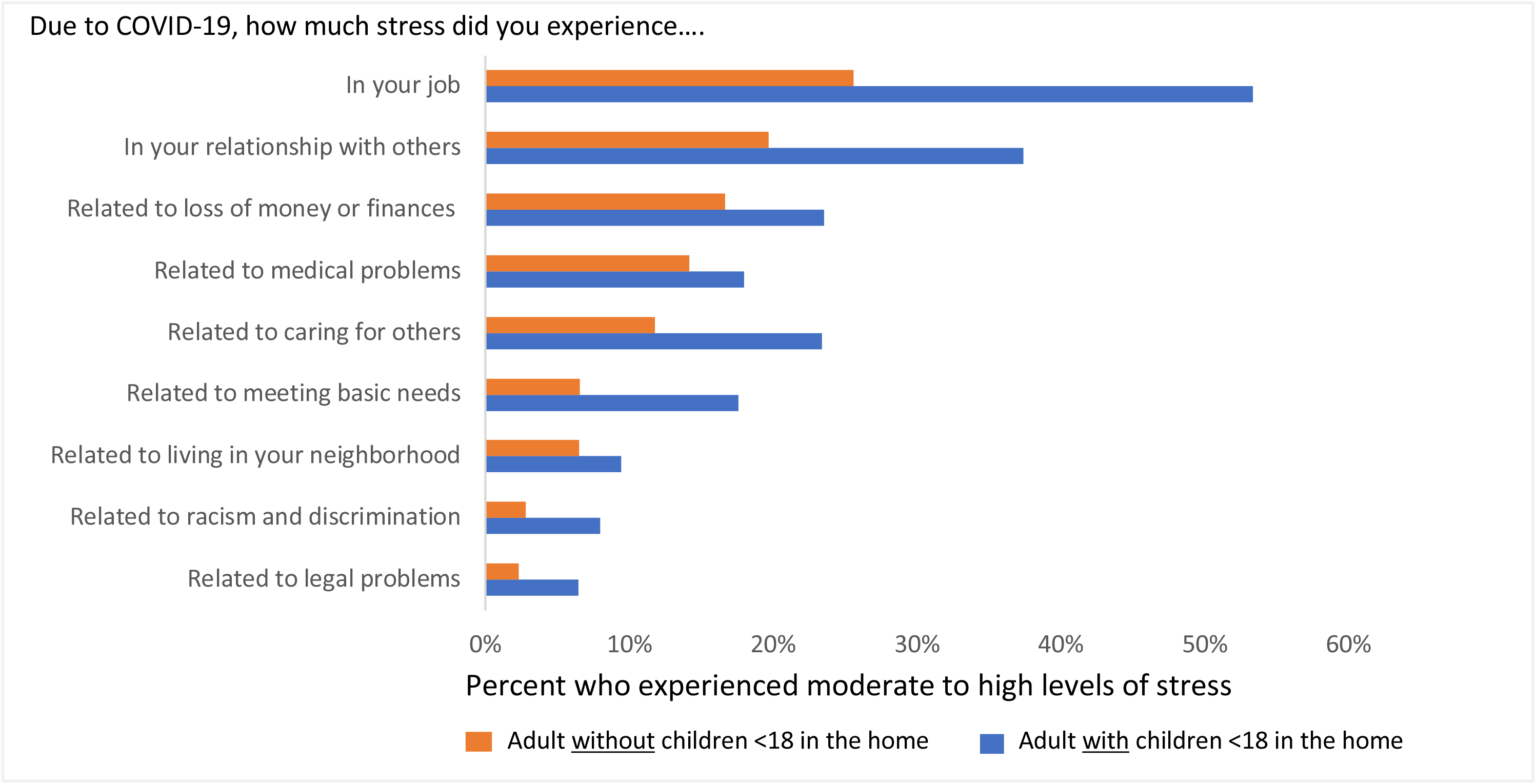
Percent of adults who self-reported experiencing moderate to high levels of stress in the following areas of their life due to COVID-19 (n=1376), comparing adults with children <18 years of age in the home to adults without children <18 years of age in the home.

## DISCUSSION

Population-specific data provide essential insights for policy makers, public health researchers and clinicians to learn and identify key factors driving risks in the population. State-wide data also offer unique opportunities to identify vulnerable populations at greatest risk of long-term impacts imposed by indicators of economic stability, social and community factors, educational opportunities, housing and family composition. These factors in turn shape access to routine medical care, can mediate or mitigate mental health and well-being, lifestyle and other opportunities for health promotion. Identification of early impacts of stay-at-home orders, closing of non-essential businesses and changes to education have shifted daily living in ways we will not understand for years to come. Early baseline information on the impact of these shifts provides and important foundation for future pandemic response and may explain differences in longer term impacts on health and well-being in communities across the United States. The goals of this initial summary of the COVID-19 impact survey results are to provide a few examples of the interconnectedness between social determinants of health and their influence in shaping how the pandemic is affecting daily lives. It is anticipated these descriptive data are an early snapshot and the data will be analyzed in greater depth by other investigators. Given space and time limitations, this is an early look and provides an overview of methods used to collect information from this unique population- based sample of adults residing in diverse geographic areas socio-economic strata.

To date, several national-level surveys have been conducted, but few statewide population-based surveys exist. Statewide data are important to monitor the impacts of the pandemic on key indicators of health and well-being in the broader context of social determinants of health. Population-specific data provide essential insights for policy makers, public health researchers and clinicians to learn and identify key factors driving risks in the population. COVID-19 short-term risks of infection, hospitalizations and death are shaped by public perceptions of risk and population adherence to public health guidelines. Results presented offer a brief description of the numerous domains within the social determinants of health impacted by the COVID-19 pandemic. Other COVID-19 surveys and public health surveillance systems have largely focused on early testing and mitigation with fewer studies aimed at tracking population health determinants and other indicators longer term. Most significantly, our findings are consistent with other national surveys and anecdotal information that females more than males are economically impacted by the COVID-19 pandemic during its early stages. Findings from this survey, conducted in the first few months (May-June) of the COVID-19 pandemic, suggest geographical location, gender, education and family dynamics are all important aspects in shaping opportunities for COVID-19 testing, access to care, mitigation efforts and coping. Early efforts to curb the public health impact of the COVID-19 pandemic were largely aimed at reducing hospitalizations, mortality and unknown consequences of longer-term morbidity from COVID-19 infections in the population. At the same time, the virus has had other serious population health consequences by impacting every aspect of our daily lives from financial well-being, to social interaction, access to care and coping mechanisms including alcohol consumption, physical activity and dietary habits. All of these social determinants of health are critical to maintain the overall health and well-being in our population. These impacts are likely to have a larger impact both positive and negative on the well-being of population health for generations to come.

### COVID-19 exposure and testing by race

Race and ethnicity are only one metric that can be used to examine impacts of SDOH on testing in the population. Given the diversity of the state population, and documented differences in the disproportionate impact of cases, hospitalizations and mortality in communities of color, we aimed to consider how these factors were shaped in this statewide population. Stark differences in testing and exposure to COVID-19 by race were seen in Wisconsin. Compared to Whites, Non-whites were 2-3 times more likely to think they had COVID-19, have been tested for COVID-19, and told they may have in contact with someone with COVID-19. These findings are consistent with those seen among n=5,834,543 individuals receiving care at US Veteran Affairs facilities between February and July, 2020, where Blacks were twice as likely to be tested for COVID-19 compared to Whites (Rentsch, et al., 2020). Indeed, rates of COVID-19 cases in the early months of the pandemic were greater among Black and Hispanics, compared to Whites (Price-Haywood, et al., 2020; Sze, et al. 2020). This was seen in the urban areas of Wisconsin, such as Milwaukee and Madison, as well as other cities across the U.S. (Ogedegbe, et al., 2020; Hooper et al., 2020). There are several potential explanations for this. First, testing was more available in urban areas in the early months of the pandemic, where health care facilities and resources are. Racial/ethnic minorities disproportionally reside in urban centers, compared to their white counterparts, where living in crowded conditions at the neighborhood and household level is more common (Ogedegbe, et al., 2020). It has also been found that racial/ethnic minorities have a disproportionate burden of underlying conditions, barriers to healthcare access, and more likely to work in essential jobs thereby increasing their risk of exposure (Ogedegbe, et al., 2020;Ruprecht, et al., 2020; Muñoz-Price et al., 2020).

### Perceived Efficacy and Behaviors related to COVID-19 Exposure Risks by Education

Sub-national population-based surveys can be used to improve understanding of the differences in trends regarding uptake of mitigation strategies and other impacts on health care access that are governed by existing social and political factors within states (Chernozhukov, et al., 2020). A study published in June of 2020, simultaneous to when this survey was administered found mandating mask wearing within states reduced growth rates in COVID-19 infections between one to two percentage points and that growth varied as states varied their implementation of mask mandates. The majority of states in this study were in the Northeast and New England (Lyu et al, 2020). A similar study by Gostin et al, found that as of July 27, 2020, thirty-one states had implemented mask mandates, but the details of the mandates- who they covered and why varied by state (Gostin, 2020). As of Feburary 10, 2021, despite federal laws requiring mask mandates in public spaced only 36 states had mask mandates in place (Lyu, 2020; Markowitz, 2021). In Wisconsin, public health mask mandates and rulings to close public gatherings were overturned by the legislature in May-June but were implemented again for public spaces in February. In May-June we found uptake of mitigation strategies and activities to cope with COVID-19 reported as fairly high. Approximately 84% of survey participants viewed face mask as effective mitigation strategy and 88% said they wore face mask, this estimate is lower compared to other countries like China where compliance with face mask usage is reported as 98% (Zhong et al., 2020). While lower than China, adherence to and reports of efficacy of mask wearing were relatively high in the study population compared to Canada and Europe were approximately 77% of participants in population surveys agreed that wearing a face mask can help prevent spread of the virus (Leigh, 2020). Much of these differences could be attributable to cultural factors and differences in messaging and risk perceptions.

A large majority of the population reported adhering to public health guidelines for mitigation early on in the pandemic, but differences by education for some key indicators did exist. Not surprisingly, education was a strong predictor of overall health status and health behaviors. At the same time our findings would suggest that key messages regarding mitigation were equally adhered to in the entire population in early May and June. Follow-up is warranted as these behaviors likely changed as the pandemic wore on and as a result of “pandemic fatigue.” Pandemic fatigue was not a key issue early on as initial shock of the COVID-19 pandemic was just beginning to take over.

### Changes in employment by gender

Among the most striking findings from this population survey, consistent with national and international trends and survey data, are apparent differences in COVID-19 effects on employment by gender. Females were more likely than males to experience disruptions in their employment, and more likely to experience a partial or full deduction of paid wages or hours in the workforce. These findings mirror those from the Current Population Survey, and the Bureau of Labor Statistics, which not only found women were more likely to experience reduced hours and wages, but more likely to leave the workforce due to the increased burden of household and childcare responsibilities (Landivar et al., 2020; Groschen et al., 2020). The Current Population Survey found the gender gap in hours worked per week from February to April 2020 grew from 4.9 to 6.2 more hours men worked compared to women (Collins et al., 2020). The patterns suggest that the COVID-19 pandemic is worsening existing gender gaps in the labor force across the U.S., and Wisconsin is no exception.

### Healthcare access and delays in care by urbanicity

Health care access and delays in care were experienced by Wisconsin residents in the early months of the pandemic. While 56% did not delay getting medical care, 44% reported delaying medical care. This aligns with a nationwide web-based survey administered to just under 5,000 adults across the U.S. June 24-30, 2020, where 40.9% avoided medical during the pandemic because of concerns about COVID-19 (Czeisler et al., 2020). Only slight differences in delays in medical care were seen by urbanicity, with urban dwellers experiencing more delays in care. While generally rural health disparities in access-to-care are a real concern, rural areas of Wisconsin did not have a surge in COVID-19 cases that mirrored the infection rates in urban areas at the time of the survey. This may explain the counter-intuitive findings, but importantly support the downstream impacts on health care access and delays in preventive care that go along with a highly infectious and deadly virus such as COVID-19.

### Health behaviors, stress and mental well-being by gender and family dynamics

Our results found COVID-19 was not only more likely to impact female’s employment, but also more likely to impact their health behaviors when compared to their male counterparts. Females were more likely to report an increase or decrease in physical activity and alcohol consumption since COVID-19, whereas males were more likely to report the same amount of each. Similar findings were seen in from a web-based convenience sample of U.S adults, where adult females were 1.28 (CI: 1.00-1.64) times more likely to report a decrease in physical activity, and also 1.47 (CI: 1.12-1.93) more likely to report an increase in physical activity, when compared with males (Knell et al., 2020). We also found women were more likely to report doing a variety of activities to cope with COVID-19, more so than males. These findings suggest COVID-19 alters the life of the females, more so than the male, from employment changes, to health behaviors, to coping strategies.

Early results of COVID-19’s impact on population health shows glimpses of the longer term impacts it will have on overall well-being, both amongst individuals and within families. Our survey found adults with children in the home were more likely to self-report moderate to high levels of stress. These results are consistent with global online survey spanning 27 countries, which found individuals with children not only reported increased stress levels compared to adults living alone or with adults, but the levels of perceived stress increased with the number of children at home (Kowal et al., 2020). The increase in stress is likely due to increased strain on childcare and remote education. Concerns over a child being sick with COVID-19 have also been found to be potential trigger of increased stress and anxiety among parents (Wang et al., 2020). Our results are consistent with a similar population-based study from Canada, where 39% of people reported worse mental/emotional health compared to before COVID-19, 26% reported worse physical health, and 31% reported worse economic health (Leigh et al., 2020). The role of child and family stress may also be impacting these trends across the population.

### Strengths and Limitations

The SHOW COVID-19 survey results offer a breadth of well characterized data to capture ongoing impacts of the COVID-19 pandemic in the population. A key strength is the breadth of focus on SDOH, its grounding in a previous statewide program. The data collected provide an important resource and baseline for tracking impacts of COVID-19 in the population well into the future. At same, it is not without limitations. This survey was administered in the early few months of the pandemic and the SHOW program had never used an online only format to interact with study participants. Response rates were thus consistent (approximately 25%) with other online surveys. At the same time the respondents were biased towards female gender and older adults. Participants were also more likely to be non-Hispanic white, have a higher education, and higher household income and better baseline health status compared to non-respondents, thus, some results may be biased to those with more time to complete the survey during the early pandemic. We aim to address these gaps in survey respondents in subsequent waves of the survey. Further, some participants across Wisconsin reported not wanting to participate in COVID-19 specific research and were willing to be in further research not focused on the pandemic. While somewhat anecdotal, there is some potential that responses are limited to those individuals who were more likely to acknowledge the pandemic as a threat to public health and thus more willing to respond. The cross-sectional nature of this first survey also limits causal interpretation, however, one could also argue that the data collected are capturing the impacts of the ongoing natural experiment that is the COVID-19 pandemic. The online format may have significant response bias; however, standardized and validated questions were used to the extent possible to improve estimates of rigor and reproducibility within the study.

### Future Directions

Moving forward, there is much to learn from the COVID-19 study and population impacts. State-specific responses and how they shape the complex interactions between SDOH and health within populations will be an important area of research to continue to follow. The COVID-19 pandemic will likely remain a threat to population health and well-being well beyond its initial economic shocks and unprecedented death toll. This study provides an early look with the potential for long-term follow-up to gain important insights on how COVID-19 is shaping the many multiple facets of health and well-being of populations. Undoubtedly, the fall-out from this pandemic will shape how lives are lived and policies to help to prepare and protect the public’s health well into the future.

## Data Availability

All data in the manuscript are available in a public data set: www.show.wisc.edu
Any data linkages with past survey waves may be requested from the data team.

https://show.wisc.edu/covid-19/covid-19-projects/covid-19-public-use-data/

## Acknowledgements

Funding for the Survey of the Health of Wisconsin (SHOW) was provided by the Wisconsin Partnership Program PERC Award (233 AAG9971). The authors would also like to thank the following investigators across several departments at the University of Wisconsin for the contributions on survey development: Janean Dilworth-Bart, Kristin Litzelman, Heather Kirkorian, Margaret Kerr, Tiffany Green, Natasha Merten, Marietou Ouayogode, Deborah Ehrenthal, Rebecca Myerson, Lydia Ashton, Thomas Oliver, Maureen Smith, Amy Trentham-Dietz, Maureen Durkin, Jane Mahoney, Marguerite Burns, and Jonathan Patz. Additional gratitude extends to the University of Wisconsin Survey Center, SHOW administrative, field, and scientific staff, as well as all the SHOW participants for their contributions to this study.

## Notes

### Competing Interest Statement

The authors have declared no competing interest.

### Clinical Protocols

http://www.show.wisc.edu

### Author Declarations

This research was approved by the University of Wisconsin Health Sciences Institutional Review Board.

### Summary of Updates

Specifics on eligible sample and final numbers of participants changed from 5502 to 5510. A secondary response rate was added and edits to final data tables to reflect final revised numbers were included as well.

