## Supplemental Materials for "Statewide Impact of COVID-19 on Social Determinants of Health - A First Look: Findings from the Survey of the Health of Wisconsin"

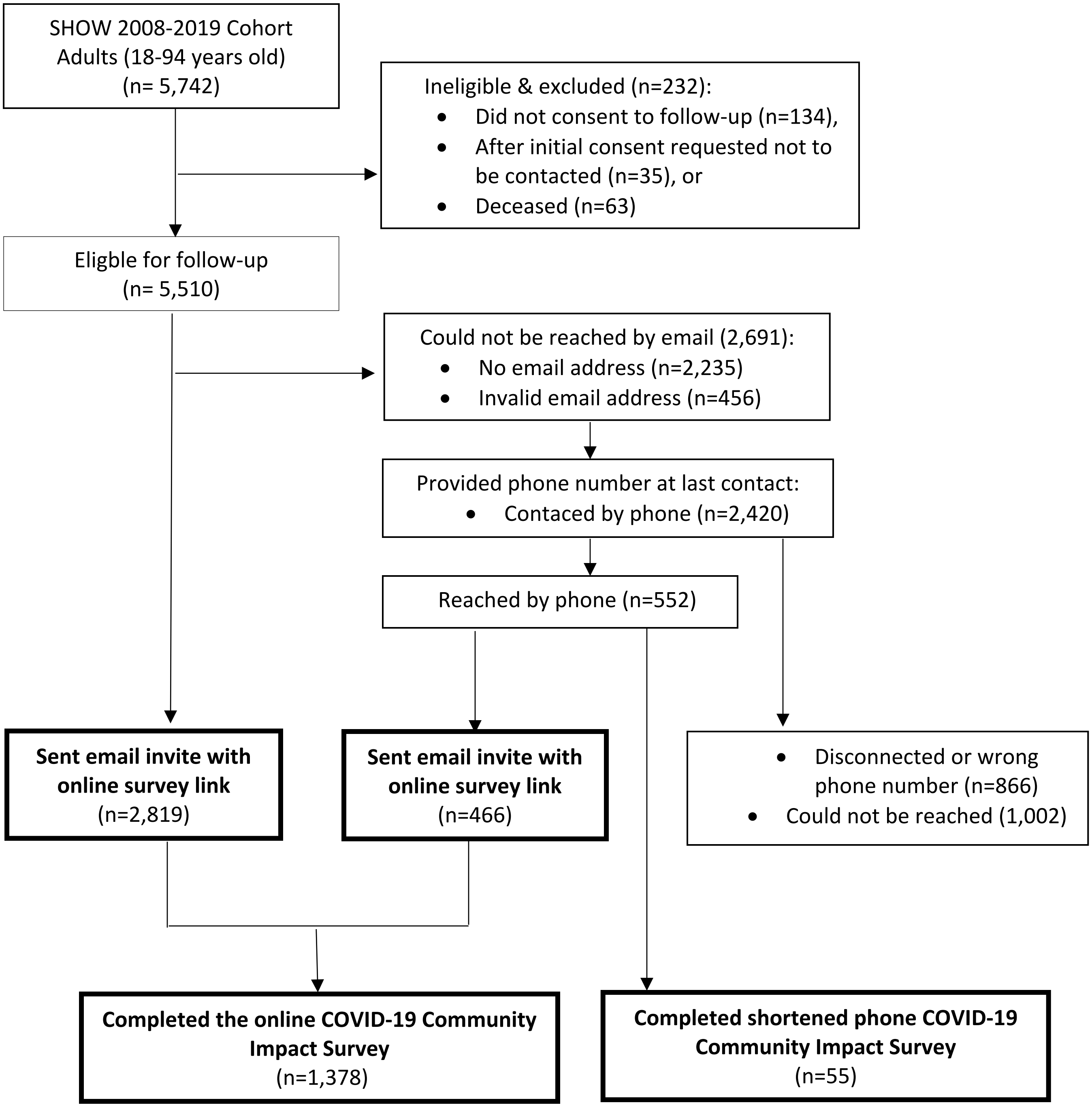


Figure 1. Flow chart of the study sample, depicting exclusion criteria and sample size.

**Supplementary Table 1. Characteristics of the study population comparing respondents to non-respondents.**

| | Total Eligible Sample (n=5510) | Respondents (n = 1378) | Non-respondents (n = 4132) | |
|---|---|---|---|---|
| | N | % | % | p-trend |
| **Gender** | | | | <.0001 |
| Male | 2398 | 36.5 | 45.9 | |
| Female | 3112 | 63.5 | 54.1 | |
| **Current Age (in years)** | | | | 0.2527 |
| 18-39 | 1108 | 18.7 | 20.6 | |
| 40-59 | 1940 | 35.3 | 35.2 | |
| 60+ | 2462 | 46.1 | 44.2 | |
| **Race** | | | | <.0001 |
| White (non-Hispanic) | 4344 | 86.7 | 76.4 | |
| AA (non-Hispanic) | 700 | 7.6 | 14.4 | |
| Hispanic | 193 | 2.3 | 3.9 | |
| Other (non-Hispanic) | 266 | 3.4 | 5.3 | |
| **Education** | | | | <.0001 |
| H.S./GED or less | 465 | 2.3 | 10.5 | |
| Some college | 3240 | 48.8 | 62.1 | |
| Bachelors or higher | 1805 | 48.9 | 27.4 | |
| **Household Income** | | | | <.0001 |
| < $25,000 | 1275 | 12.8 | 28.0 | |
| $25,000 - $49,999 | 1262 | 22.2 | 24.4 | |
| $50,000 - $99,999 | 1726 | 35.5 | 31.7 | |
| >$99,999 | 1024 | 29.5 | 15.9 | |
| **Household size** | | | | <.0001 |
| 1 | 1125 | 15.2 | 23.0 | |
| 2 | 2130 | 46.1 | 37.6 | |
| 3-4 | 1515 | 29.5 | 27.9 | |
| 5+ | 590 | 9.3 | 11.6 | |
| **Urbanicity** | | | | 0.3639 |
| Urban | 3724 | 68.6 | 67.3 | |
| Rural | 1786 | 31.4 | 32.7 | |
| **Self-reported Health** | | | | <.0001 |
| Excellent | 454 | 11.9 | 8.6 | |
| Very Good | 1920 | 47.1 | 37.7 | |
| Good | 1768 | 32.6 | 38.8 | |
| Fair | 525 | 7.3 | 12.4 | |
| Poor | 104 | 1.1 | 2.6 | |
| **Chronic Health Condition** | | | | |
| Asthma and/or COPD | 673 | 11.5 | 12.5 | 0.3273 |
| Cardiovascular disease | 1977 | 34.4 | 36.4 | 0.1852 |
| Diabetes (any type) | 593 | 8.2 | 11.6 | 0.0004 |

AA: African American; N: number; H.S.: high school; GED: General Education Development test; COPD: Chronic Obstructive Pulmonary Disease
p-trend: statistically significany difference of demographics by response to COVID-19 survey using Chi-square test
Urbanicity: considered rural if residence located in rural census block group defined by the U.S. Census Bureau as having fewer than 2,500 people
Cardiovascular disease: if reported history of heart failure, heart attack, angina, stroke, high cholesterol, or transient ischemic attack
Asthma/COPD: self-reported current asthma, asthma episode(s) in last 12 months, or current asthma treatment

**Supplementary Table 2.** Percentage of respondents who have done the following mitigation behaviors by race.

| | Survey Sample (n = 1378) n (%) | White (n=1194) n (%) | Non-White (n=182) n (%) | P-trend |
|---|---|---|---|---|
| **Do you think you may have had COVID-19 at any time since COVID-19 began?** | | | | 0.0003 |
| Yes | 128 (11.0) | 97 (9.6) | 31 (20) | |
| No | 1036 (89.0) | 912 (90.4) | 124 (80) | |
| **Have you been told by a health care professional that you have or had COVID-19?** | | | | 0.022 |
| Yes | 9 (1) | 5 (0.42) | 4 (2.2) | |
| No | 1361 (99) | 1183 (99.6) | 178 (97.8) | |
| **Have you ever been tested for COVID-19?** | | | | <0.0001 |
| Yes | 94 (6.8) | 68 (5.7) | 26 (14.2) | |
| No | 1280 (93.2) | 1123 (94.3) | 157 (85.8) | |
| **What were the test results?** | | | | 0.58 |
| Positive | 4 (4.3) | 2 (2.9) | 2 (7.7) | |
| Negative | 70 (74.5) | 51 (75) | 19 (73) | |
| Still waiting for results | 20 (21.3) | 15 (22.1) | 5 (19.2) | |
| **Did you try to get tested but were turned away?** | | | | 0.016 |
| Yes | 28 (2.2) | 20 (1.8) | 8 (5.1) | |
| No | 1248 (97.8) | 1099 (98.2) | 149 (94.9) | |
| **Contacted by health professional about potential exposure to COVID-19** | | | | <0.0001 |
| Yes | 50 (3.7) | 33 (2.8) | 17 (9.6) | |
| No | 1310 (96.3) | 1149 (97.2) | 161 (90.4) | |
| **If you were diagnosed with COVID-19, or believe you had COVID-19, were you afraid or embarrassed to disclose this information to your friends or employer?** | | | | <0.0001 |
| Yes | 32 (2.3) | 22 (1.9) | 10 (5.7) | |
| No | 617 (45.7) | 518 (44.1) | 99 (55.9) | |
| N/A | 702 (52.0) | 634 (54) | 68 (38.4) | |
| **During the COVID-19 outbreak, do or did you experience stigma or discrimination from other people because of your identity, having symptoms or other factors related to COVID-19?** | | | | 0.005 |
| Yes | 80 (6.3) | 61 (5.5) | 19 (11.5) | |
| No | 1192 (93.7) | 1045 (94.5) | 147 (88.5) | |

Column percents shown.
p-trend: statistically significant difference of demographics by race using Chi-square test

**Supplementary Table 3.** (a) Percentage of respondents who perceive the following as "Somewhat effective or "Very effective" at keeping them safe from COVID-19. (b) Percentage of respondents who self-reported having done the following mitigation behaviors by highest education level attained.

| **(a) How effective are the following actions are at preventing you from getting COVID-19?** *(select all that apply)* | **Survey Sample (n = 1378) %** | **< = H.S. / G.E.D/ Some College (n=680) %** | **Bachelor's, Graduate or Professional (n=697) %** |
|---|---|---|---|
| Doing nothing | 17% | 21% | 12% |
| Avoiding outside exercise | 19% | 23% | 15% |
| Praying | 48% | 57% | 39% |
| Getting tested for COVID19 | 53% | 49% | 56% |
| Wearing a facemask | 84% | 84% | 85% |
| Social distancing | 94% | 93% | 94% |
| Avoiding public spaces & gatherings | 94% | 93% | 96% |
| Washing hands | 98% | 98% | 99% |
| **(b) Have you done any of the following because of COVID-19?** *(select all that apply)* | **Survey Sample (n = 1378)** | **< = H.S. / G.E.D/ Some College (n=680)** | **Bachelor's, Graduate or Professional (n=697)** |
| Doing nothing | 17% | 21% | 12% |
| Avoiding outside exercise | 19% | 23% | 15% |
| Praying | 48% | 57% | 39% |
| Getting tested for COVID19 | 53% | 49% | 56% |
| Wearing a facemask | 84% | 84% | 85% |
| Social distancing | 94% | 93% | 94% |
| Avoiding public spaces & gatherings | 94% | 93% | 96% |
| Washing hands | 98% | 98% | 99% |
| Self-quarantined | 32% | 29% | 33% |
| Stayed at home most of the time | 89% | 83% | 91% |
| Practiced social distancing (6 ft) | 93% | 88% | 95% |
| Routinely washed hands (>= 20 secs.) | 88% | 84% | 89% |
| Worn a mask | 89% | 86% | 90% |
| Avoided shaking hands | 89% | 83% | 91% |
| Visited elderly relatives | 10% | 9% | 10% |
| Visited friends or relatives | 23% | 22% | 23% |
| Bought food for elderly relatives | 14% | 13% | 13% |
| Bought food for friends and family | 17% | 17% | 17% |
| Taken your temperature regularly | 20% | 23% | 16% |
| Took public transportation to work | 1% | 1% | 0% |
| Cancelled a social gathering | 57% | 50% | 63% |
| Cancelled travel plans | 54% | 43% | 62% |
| None of these | 1% | 0% | 1% |
| Other | 4% | 2% | 5% |

**Supplementary Table 4.** Current employment status (n=1378) by employment or school status prior to COVID-19.

| What is your job or employment status right now? (May/June 2020) | Total (n=1378) n (%) | Did you have a paid job and/or were you in school before COVID-19? Yes (n=821) n (%) | No (n=556) n (%) |
|---|---|---|---|
| Working full-time for pay | 54[illegible] (39) | 519 (96) | 24 (4) |
| Retired and not looking for a job | 463 (33) | 31 (7) | 432 (93) |
| Working part-time for pay | 150 (11) | 144 (96) | 6 (4) |
| Other | 98 (7) | 62 (63) | 36 (37) |
| Not working for pay and not looking for a job | 85 (6) | 39 (46) | 46 (54) |
| Not working for pay and looking for a job | 39 (2.8) | 25 (64) | 14 (35.9) |
| Student | 13 (0.9) | 10 (77) | 3 (23) |

**Supplementary Table 5.** (a) Percent who delayed getting care for the following reasons (n= 1378), (b) Among those who experienced delay in care (n=603), the percent who reported the type of appointments and/or procedures missed. (c) Percent who indicated there was a time when they needed the following but could not get it because of COVID-19 (n=1378), (d) The percent who received telemedicine or telehealth since COVDI-19 began (n=1378). Participant is considered to reside in rural location if residence is located in rural census block group defined by the U.S. Census Bureau as having fewer than 2,500 people.

| (a) **Have you delayed getting care of any of the following reasons due to COVID-19?** *(select all that apply)* | **Survey Sample (n = 1378) %** | **Urban (n=945) %** | **Rural (n=433) %** |
|---|---|---|---|
| No Delay in care | 56% | 54.6% | 59.1% |
| Postponed or cancelled due to COVID-19 | 34% | 34.4% | 34.4% |
| Afraid to get care because of COVID-19 | 8.9% | 8.8% | 9.0% |
| Could not get an appointment soon enough | 5.1% | 5.0% | 5.3% |
| Clinic/office not open when arrived | 2.4% | 2.5% | 2.1% |
| Could not get through on the telephone | 1.2% | 1.2% | 1.4% |
| Did not have transportation | 0.7% | 0.9% | 0.2% |
| Went, but wait was too long | 0.6% | 0.7% | 0.2% |
| Other | 2.5% | 2.7% | 2.3% |
| Don't know | 1.9% | 2.2% | 1.2% |
| (b) *Those who experienced delay in care,* **Which of the following types of healthcare appointments were delayed or canceled due to the COVID-19?** *(select all that apply)* | **Survey Sample (n = 603) %** | **Urban (n=429) %** | **Rural (n=177) %** |
| Eye doc/optometrist | 18% | 19% | 17% |
| Mammogram[a] | 18% | 14% | 27% |
| Blood draw | 11% | 10% | 14% |
| Pap smear/cervical cancer screening[a] | 7% | 7% | 8% |
| Skin mole/skin cancer screening | 6% | 7% | 6% |
| PT/occupational Therapy | 6% | 7% | 5% |
| Colonoscopy | 5% | 3% | 7% |
| Chiropractor | 5% | 6% | 3% |
| Allergy | 2% | 2% | 2% |
| Asthma or COPD | 1% | 2% | 1% |
| Cardiac rehab | 0% | 1% | 0% |
| Lung cancer screening/CT/chest x-ray | 0% | 1% | 0% |
| (c) **Was there any time when you needed any of the following but could not get it becuase of COVID-19?** *(select all that apply)* | **Survey Sample (n = 1378) %** | **Urban (n=945) %** | **Rural (n=433) %** |
| Regular Health Care | 22% | 23% | 20% |
| Dental Care | 35% | 34% | 36% |
| Eyeglasses | 13% | 13% | 12% |
| Mental Health care | 3% | 3% | 2% |
| Prescription Medication | 3% | 3% | 2% |

| | | | |
|---|---|---|---|
| None of These | 49% | 48% | 52% |
| Don't know | 1% | 1% | 0% |
| **(d) Since COVID-19 began, did you receive telemedicine or telehealth?** | **Survey Sample (n = 1378) %** | **Urban (n=945) %** | **Rural (n=433) %** |
| Yes | 22 | 22 | 21 |
| No | 78 | 78 | 79 |

[a]Only asked among those self-identified as female and reported a delay in care due to COVID-19 (n=432)

**Supplementary Table 6. The impact of COVID-19 on health-related behaviors by gender[a]** (a-c) Respondents reported the degree to which their physical activity level, alcohol consumption and smoking quantity changed since COVID-19 began, compared to before COVID-19. Only current alcohol consumers (n= 1285) and smokers (n=115) were asked about changes in their alcohol and smoking quantity. (d) Percentage of respondents who reported doing the following activities to help cope with stress from COVID-19. (a-d) n=5 nonconforming/transgender were excluded because too few in self-identied gender category to look at separately in this analyses.

| (a) How has your overall level of physical activity changed due to COVID-19? | Survey Sample (n = 1367) % | Male (n=502) % | Female (n=865) % |
|---|---|---|---|
| A lot more | 17% | 13% | 20% |
| A little more | 27% | 26% | 27% |
| About the same | 33% | 41% | 29% |
| A little lower | 16% | 14% | 17% |
| Much lower | 6% | 5% | 7% |
| **(b) In the last 60 days during COVID-19, would you say the amount of alcohol you drink now compared to before is:** | **Survey Sample (n = 1285) %** | **Male (n=468) %** | **Female (n=817) %** |
| A lot more | 5% | 4% | 6% |
| A little more | 18% | 16% | 19% |
| About the same | 61% | 65% | 59% |
| A little lower | 7% | 6% | 7% |
| Much lower | 9% | 9% | 9% |
| **(c) In the last 60 days during COVID-19, how would you comnpare your smoking/vaping now to before COVID-19:** | **Survey Sample (n = 115) %** | **Male (n=38) %** | **Female (n=77) %** |
| A lot more | 14% | 8% | 16% |
| A little more | 30% | 24% | 34% |
| About the same | 47% | 61% | 42% |
| A little lower | 5% | 5% | 5% |
| Much lower | 3% | 3% | 4% |
| **(d) Since COVID-19, have you done any of the following as a way to cope with COVID-19?** *(select all that apply)* | **Survey Sample (n = 1369) %** | **Male (n=497) %** | **Female (n=872) %** |
| None of these | 3% | 4% | 2% |
| Wrote in a journal | 8% | 5% | 11% |
| Meditating | 11% | 7% | 14% |
| Yoga | 12% | 5% | 15% |
| Worked on art projects | 23% | 8% | 31% |
| Attended online religious/spiritual event | 34% | 27% | 37% |
| Prayer | 39% | 27% | 46% |
| Exercised/worked out | 43% | 42% | 44% |
| Played music | 48% | 38% | 53% |
| Read books for pleasure | 49% | 36% | 57% |
| Played games (online, card, board) | 53% | 46% | 57% |
| Baked and cooked | 60% | 39% | 72% |

| | | | |
|---|---|---|---|
| Video call with friends/family | 61% | 51% | 66% |
| Watched online movies/shows | 63% | 61% | 64% |
| Gardened/home improvement projects | 64% | 60% | 66% |
| Took walks outside | 71% | 66% | 74% |
| Watched television | 79% | 75% | 81% |

**Supplementary Table 7a. Children in the home and caregiving during COVID-19.** Percent of adults who report having children in the home, caregiving for children and/or adults during COVID-19.

| **Have one or more children (< 18 years of age) living in your home:** | **Survey Sample (n = 1359) N (%)** |
|---|---|
| Yes | 404 (30) |
| No | 955 (70) |
| *Among those with children <18 years of age in the home,* **Do you currently care for any of the children in your household during the COVID-19 pandemic?** | **(n = 401) N %** |
| Yes | 352 (88) |
| No | 49 (12) |
| **Are you currently providing care for an adult (18+ years of age) in your household that has an illness or a disability?** | **(n = 1158) %** |
| Yes | 70 (6) |
| No | 1088 (94) |

**Supplementary Table 7b. Stress during COVID-19 by the status of children in the home.** Percent of adults who self-reported experiencing moderate to high levels of stress in the following areas of their life due to COVID-19, comparing adults with children <18 years of age in the home to adults without children <18 years of age in the home.

| **Due to COVID-19, how much stress did you experience…** *(select all that apply)* | Percent who self-reported experiencing moderate to high level of stress | | |
|---|---|---|---|
| | **Survey Sample (n = 1378)** | **Adults with Children[a] in the home (n=404)** | **Adults without Children[a] in the home (n=473)** |
| Related to legal problems | 4% | 6% | 2% |
| Related to racism and discrimination | 4% | 8% | 3% |
| Related to living in your neighborhood | 7% | 9% | 7% |
| Related to meeting basic needs | 10% | 18% | 7% |
| Related to caring for others | 15% | 23% | 12% |
| Related to medical problems | 16% | 18% | 14% |
| Related to loss of money or finances | 19% | 24% | 17% |
| In your relationship with others | 25% | 37% | 20% |
| In your job | 34% | 53% | 26% |

[a]Children includes anyone < 18 years of age
